# SARS-CoV-2 genomic characterization and clinical manifestation of the COVID-19 outbreak in Uruguay

**DOI:** 10.1101/2020.10.08.20208546

**Authors:** Victoria Elizondo, Gordon W. Harkins, Batsirai Mabvakure, Sabine Smidt, Paul Zappile, Christian Marier, Matthew Maurano, Victoria Perez, Natalia Mazza, Carolina Beloso, Silvana Ifran, Mariana Fernandez, Andrea Santini, Veronica Perez, Veronica Estevez, Matilde Nin, Gonzalo Manrique, Leticia Perez, Fabiana Ross, Susana Boschi, Maria Noel Zubillaga, Raquel Balleste, Simon Dellicour, Adriana Heguy, Ralf Duerr

## Abstract

COVID-19 is a respiratory illness caused by severe acute respiratory syndrome coronavirus 2 (SARS-CoV-2) and declared by the World Health Organization a global public health emergency. Among the severe outbreaks across South America, Uruguay has become known for curtailing SARS-CoV-2 exceptionally well. To understand the SARS-CoV-2 introductions, local transmissions, and associations with genomic and clinical parameters in Uruguay, we sequenced the viral genomes of 44 outpatients and inpatients in a private healthcare system in its capital, Montevideo, from March to May 2020. We performed a phylogeographic analysis using sequences from our cohort and other studies that indicate a minimum of 23 independent introductions into Uruguay, resulting in five major transmission clusters. Our data suggest that most introductions resulting in chains of transmission originate from other South American countries, with the earliest seeding of the virus in late February 2020, weeks before the borders were closed to all non-citizens and a partial lockdown implemented. Genetic analyses suggest a dominance of S and G clades (G, GH, GR) that make up >90% of the viral strains in our study. In our cohort, lethal outcome of SARS-CoV-2 infection significantly correlated with arterial hypertension, kidney failure, and ICU admission (FDR < 0.01), but not with any mutation in a structural or non-structural protein, such as the spike D614G mutation. Our study contributes genetic, phylodynamic, and clinical correlation data about the exceptionally well-curbed SARS-CoV-2 outbreak in Uruguay, which furthers the understanding of disease patterns and regional aspects of the pandemic in Latin America.

## Introduction

The novel coronavirus SARS-CoV-2, first discovered in Wuhan, China, in December 2019, rapidly extended throughout the globe, and among the >35 million confirmed positive people from >210 affected countries and territories, >1 million have died from the rapidly-spreading SARS-CoV-2 virus as of October 5th, 2020 [1].

Even though South America was mostly spared in the early months of the pandemic and was the last continent where it spread, it was severely hit with the arrival of the fall season to the Southern hemisphere. The virus is currently ravaging Latin America, with Brazil, Colombia, Argentina, Peru, and Mexico among the ten countries with the highest numbers of cases worldwide. In contrast, Uruguay, a small country located south of Brazil, has succeeded in maintaining a very low number of total cases (2,145) through the closing of its borders, a partial lockdown, and an early Test, Trace and Isolate (TETRIS) strategy [2]. Uruguay has a population of ∼3.3 million inhabitants, of which more than a third (∼1.3 million) live in the capital city of Montevideo, according to the last census in 2011 [3]. Montevideo is Uruguay’s most interconnected city due to the presence of its international airport and harbor. The first positive case was officially registered on March 13th, and almost seven months later, 245,000 samples (>7% of the total population of Uruguay) had been tested. In March 2020, after the first four positive cases of SARS-CoV-2 were reported in Montevideo, the government issued a responsible voluntary quarantine in the country, involving the closure of schools, public entities, and businesses, urging the population to stay home [4]. On March 24th, land, maritime, and air borders were closed, allowing only Uruguayan citizens to enter the country [5]. As of October 5th, 2020, 2,145 patients have tested positive, 1,831 recovered, 243 are active cases, and 48 patients died [6].

Viral genome sequencing (genomic surveillance) is a powerful approach to determine the origin of pathogen introductions into a certain location and trace and track the virus’s subsequent spread and evolution. It has been utilized to complement other epidemiological parameters determined by testing, contact tracing and implementation of other public health measures such as lockdowns [2,7-12].

Although preliminary phylogenetic analyses of Uruguayan SARS-CoV-2 sequences have previously been communicated on pre-print websites [2,7], there remains a critical lack of knowledge regarding SARS-CoV-2 mutation, dispersal, and transmission patterns, and whether statistical associations exist among viral genomic, demographic, and clinical features. For example, a detailed phylogeographic analysis has not been undertaken to investigate the global dispersal dynamics with respect to Uruguay.

In the present study, we describe and build on genomic surveillance of a private nonprofit healthcare system in Uruguay’s capital, Montevideo, where SARS-CoV-2 cases have been identified since March 17th. We have sequenced and characterized 44 positive cases spanning March to late May. We build on the existing Nextstrain framework, a broadly used and universally recognized analytical platform for the analysis of global SARS-CoV-2 viral genome sequence data that has accumulated since the start of the pandemic [13] to analyze newly sequenced SARS-CoV-2 genomes sampled in Uruguay, and combine it with Bayesian Evolutionary Analysis of Sampling Trees (BEAST).

An important aim of this study is to identify the different SARS-CoV-2 introduction events in Uruguay, located in a geographical region of the pandemic that has been understudied so far. Specifically, we aim at (i) identifying and investigating the importance of independent introduction events in establishing the COVID-19 epidemic in Uruguay, (ii) analyzing the spatial distribution of the resulting clades in the capital city, Montevideo, (iii) looking for phylogenetic clusters within sampled institutions (hospitals, nursing homes), iv) determining whether statistically significant correlations exist among mutations, demographic, and clinical parameters.

## Materials and Methods

### Bioethics, sample collection & RNA extraction

The molecular lab at the Asociación Española Primera en Salud (AEPS) is a fully accredited clinical lab, regulated by the Uruguayan Ministry of Health (Ministerio de Salud Púbica, MSP). Sampling was done according to the regional IRB guidelines and the recommendations of the MSP. Our study was evaluated by the commission of bioethics and ethics in research of the AEPS, headed by Fernando García [14-16]. IRB approval was waived, because the genetic analysis was restricted to the virus and not the host, and clinical correlation analyses were done on fully de-identified samples (UY ID and GISAID IDs were given by the Uruguayan and New York research teams, respectively, and are not traceable to any medical record number). The participants provided informed oral consent, and the data were analyzed anonymously. For participants <18 years of age, formal written or verbal consent was obtained from the parent/guardian at sampling, and data were kept anonymously for the entire study. The molecular clinical lab at AEPS is accredited for diagnostic RT-PCR tests for SARS-CoV-2 (COVID-19) in Uruguay. Naso-oropharyngeal swabs were collected in viral transport media and RNA was extracted using the QIAsymphony⍰ DSP Virus/Pathogen Mini or Midi kit (Qiagen), respectively, and confirmatory qualitative commercial RT-PCR kits were used for diagnosis and screening (depending on critical availability during the outbreak) (**Table 1**). Full details in **Supplemental Methods**.

**Table 1.** Overview of three different SARS CoV2 detection kits included in this study

| Real-time RT-PCR System | Country | Regulatory Status | Target Gene(s) | Limit of Detection (copies/reaction) | Run Time (hours) | PCR Machine |
| --- | --- | --- | --- | --- | --- | --- |
| LightMix Modular SARS and Wuhan CoV E/RdRp/N-gene (TIB MOLBIOL) | Germany | RUO | E, N and RdRp | 5-10 | 1.2 | LightCycler® z480 System, ROCHE |
| GeneFinder COVID-19 Plus Real Amp Kit (Healthcare) | Korea | HealthCanada<br>BrazilANVISA<br>SingaporeHSA<br>CE-IVD | E, N and RdRp | 10 | 2.2 | CFX96 Touch Real-Time PCR Detection System, BIO-RAD |
| RealStar SARS- CoV-2 RT-PCR kit 1.0 (Altona) | Germany | US FDAEUA<br>CE-IVD | E, S | NA | 2.25 | CFX96 Touch Real-Time PCR Detection System, BIO-RAD |

### Library Preparation, Sequencing, and Read Processing

To amplify the viral genomes in preparation for sequencing, we used the Swift Normalase Amplicon Panel (SNAP) SARS-CoV-2 Panel (Swift Biosciences, Whole viral genome single tube NGS assay, cat# SN-5XCOV296). The libraries were run on an Illumina NovaSeq 6000 300 cycle flow cell, as paid end 150, using dual indices. Both positive and negative control samples were also run during library prep but not sequenced. The negative control sample was water, and the positive control sample consisted of SARS-CoV-2 genome (Twist Biosciences, cat# 102024) serially diluted to 100 viral copies mixed into 50 ng of Universal Human Reference RNA (Agilent, cat# 18091050). Sequencing reads were demultiplexed with Illumina bcl2fastq v2.20 requiring a perfect match to indexing barcode sequences, and aligned to the reference SARS-CoV-2 genome (NC_045512.2, wuhCor1). Only samples with >23,000 bp unmasked sequences were further analyzed, and variants were called using bcftools v1.9. Details of the library generation and sequencing read processing are in the **Supplemental Methods**.

### Statistics

D’Agostino & Pearson normality tests were performed to assess whether data values followed Gaussian distribution and whether parametric or nonparametric statistical tests were indicated (GraphPad Prism v.8). As a result, correlation analyses were done using nonparametric Spearman rank tests. Correlation coefficients (*r*), *P*-values, adjusted *P*-values, and *q* values were calculated in Prism. *P*-values were adjusted for multiple comparisons using Holm-Sidak (α = 0.05). The false discovery rate (FDR) of q was calculated at 0.5%, 1% and 5%. Correlation analyses with a sample size of 44 had 80.7% power (**α** = 0.05) to distinguish correlation coefficients that differ by 0.4 standard deviation units (G*Power 3.1.9.4).

### Genetic analysis

Sequence retrieval and multiple sequence alignment. SARS-CoV-2 reference sequences were downloaded from GISAID EpiCoV and combined with our Uruguayan study sequences in MEGA v.5.2 software, also used for sequence quality analysis, capping, and data refinement, if applicable [17]. Sequence alignments were performed using MAFFT v7.471, FFT-NS-2 method [18]. Highlighter analyses were performed on MAFFT-aligned full SARS-CoV-2 sequences from our Uruguayan study cohort with reference sequence Wuhan-Hu-1 as a master using the Highlighter tool provided by the Los Alamos HIV sequence database [19].

### Inference of a time-scaled phylogeny

To infer our SARS-CoV-2 time-scaled phylogenetic tree, we selected global reference sequences used for the Nextstrain analysis specific for South America as of August 6th, 2020. This dataset consisted of 1747 sequences sampled between December 26th, 2019 and July 17th, 2020, from Africa (40), Asia (194), Europe (112), North America (45), Oceania (17), and South America (128). We added 74 SARS-CoV-2 Uruguayan sequences to generate an initial dataset containing 1821 viral whole-genome sequences (Table S2). Using the Nextstrain metadata to identify the accessions of interest, we then downloaded the latest whole-genome sequence alignment from the GISAID database.

We aligned the whole genome sequences using MAFFT v7.471 [18] and manually edited these by trimming the 5’and 3’ untranslated regions and removing any gap only sites and low-quality sequences. This resulted in one low-quality Uruguayan sequence being removed, and a total of 73 remaining. The manually-edited alignment was then used to construct a maximum likelihood tree with ultra-fast bootstraps of 1000 replicates in IQ-TREE version 2.0.3 [20], using the GTR+F+I nucleotide substitution model selected by Bayesian information criterion using model test implemented in IQ-TREE. TempEst [21] was used to check for outlier sequences in the tree resulting in the removal of a further ten sequences, to make up a final data set of 1810 sequences. The tree was dated using TreeTime version 0.7.6 [22], specifying a clock rate of 8 x 10-4 substitutions per site per year to replicate the original Nextstrain workflow analysis as faithfully as possible.

### Phylogeography analyses

To obtain an estimate of the number of independent introductions of SARS-CoV-2 into Uruguay, we performed a preliminary phylogeography analysis using the asymmetric rates discrete diffusion model implemented in BEAST version 1.10.4 [23], adopting the fixed tree approach [11]. This approach greatly reduces the computational time required to perform the analyses by annotating the phylogeography analyses onto a fixed time-scaled phylogenetic tree. Our model considered two discrete ancestral location states, i.e., Uruguay and non-Uruguay, and specified a Markov chain Monte-Carlo (MCMC) length of 1 million steps, sampling every 200 steps to produce a posterior distribution of trees containing 5,000 trees. The time-scaled maximum clade credibility tree (MCC) tree from the discrete model phylogeography analysis conducted in BEAST was then identified using TreeAnnotator, available as part of the BEAST package and visualized in FigTree v.1.4.4 [24]. The BEAST log files were inspected for convergence using Tracer version 1.7.1 [25]. All model parameters achieved effective sample size (ESS) values >200 indicating sufficient mixing and convergence to stationary.

To estimate the potential regional source(s) of the independent introductions into Uruguay, we then replicated this phylogeography analysis, but this time considered seven discrete ancestral location states, including Uruguay, Africa, South America, North America, Oceania, Asia, and Europe.

### Software Scripts and Visualization

See Supplemental Methods.

## Results

### Study population and clinical parameters

A total of 44 diagnosed positive COVID-19 participants were included in this study, 25 men and 19 women with a comparable mean age (and range) of 54 (15-92) and 59 (24-89) years, respectively. Sixty eight percent of the participants did not require hospitalization, 18% were hospitalized or received in-home care (9% on ventilation and 5% on ICU), and 14% were deceased. Whereas the early COVID-19 positive cases in March were predominantly clade S infections, clade G viruses subsequently became dominant in April and May 2020. **(Figure 1 A-C)**.

**Figure 1.**
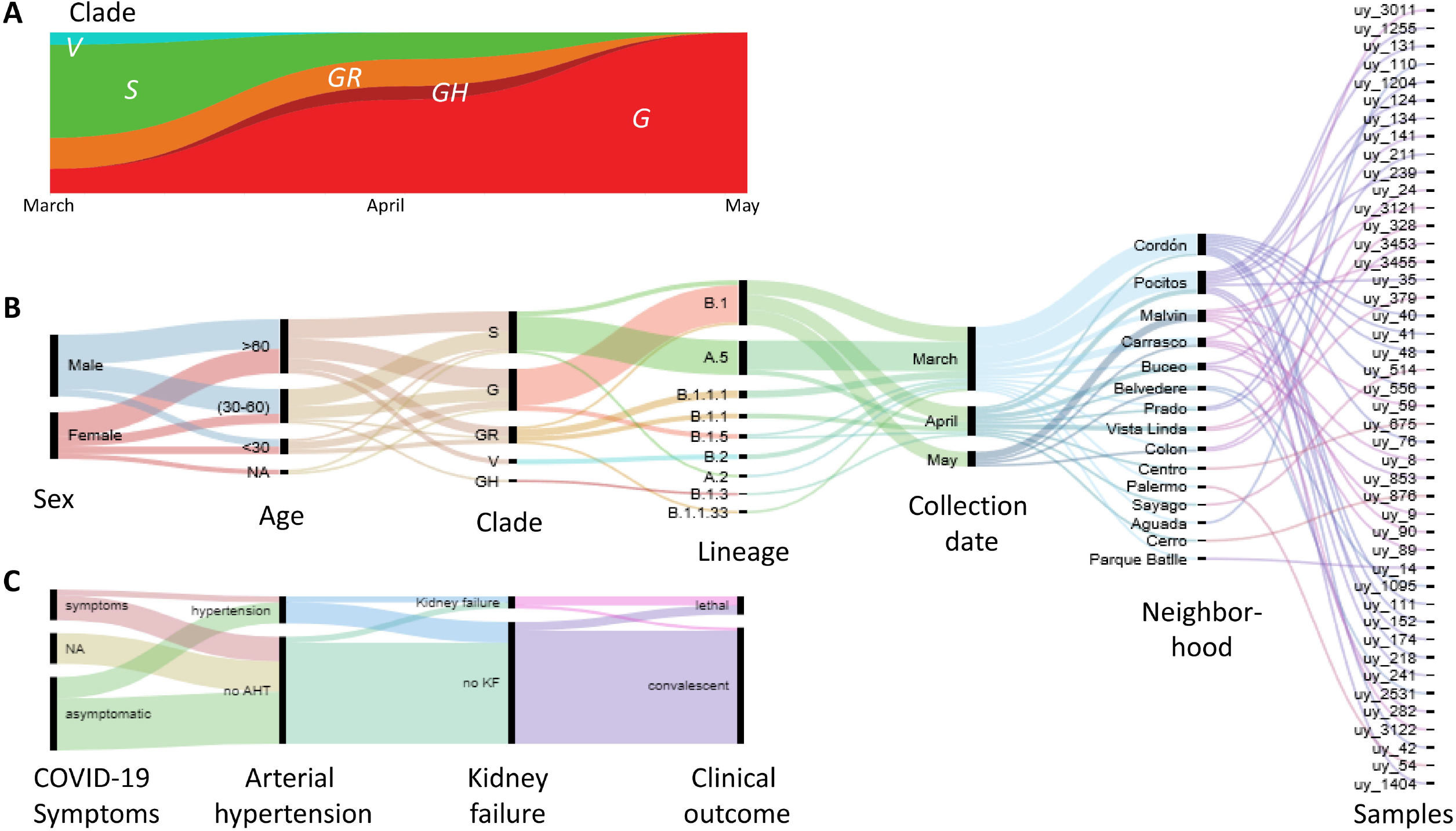
Virologic, demographic, and clinical parameters of the Uruguayan study cohort. **(A)** SARS-CoV-2 clade distribution over the ∼3 month study period from March to May. **(B)** Demographic, virologic, and sample collection data are shown in a multicategorical alluvial diagram with the display of relatedness among features of two neighboring nodes. **(C)** Clinical parameters of study participants, shown in alluvial representation as in B.

Our cohort’s cases came from 15 neighborhoods in the capital, Montevideo. The majority of the samples came from the central neighborhoods of Cordón and Pocitos, where the two biggest outbreaks within our cohort occurred (**Figure 1B and Figure S1**). Only two participants (from the Montevideo neighborhoods Pocitos and Prado) reported a possibility of travel-related infection after returning from Japan.

### Diverse SARS-CoV-2 mutation profiles with increased prevalence of spike D614G variants and associated mutations in the evolving epidemic

The SARS-CoV-2 genetic variants in our study group are very diverse and comprise sequences from clades S, V, G, GH, and GR (**Figures 1A and 2A, Table S1**). We found 446 mutations in our Uruguayan study sequences compared to the reference Wuhan.Hu.1 sequence [26], including three gaps. 313 were SNPs (non-amino acid changing), scattered across 60 positions, and 130 were amino acid-changing, scattered across 32 positions of the open reading frames (**Figure 2A, B**). We observed between 5-12 single nucleotide polymorphisms (SNP) resulting in 1-7 amino acid (AA) replacements per viral genome (**Figure 2B, Figures S2 and S3**). In chronologically sorted mutation/highlighter plots, various mutation patterns appeared in the first 2/3 of the study period, whereas in the last 1/3, the mutation patterns became more homogenous. In March and early April, ORF8 mutation L84S and the associated C8782T single nucleotide polymorphism (SNP) were most abundant (>1/3 of sequences) (**Figure 2A, C, Figures S2 and S3**). Both mutations have become known as clade S-defining mutations (mostly in sublineage A.5) [27]. In addition, we found three more SNPs to be significantly associated, namely C17470T, C25521T, and C26088T that all together build a strongly significant, positive correlation cluster composed of five mutations (**Figure S4**). Interestingly, after April, most sequences belonged to clade G (sublineage B.1), defined by the spike protein’s D614G mutation. 57% of our cohort’s sequences contain the D614G mutation co-occurring with SNPs C241T, C3037T, and C14408T, the latter causing the AA replacement P323L in the RNA-dependent RNA polymerase (RdRP) gene of nsp12 (**Figure 2A, C, Figures S2 and S4**). As known from other global studies [27], clade S and G-associated mutations are largely exclusive, resulting in strongly inverse correlation clusters (**Figure S4**).

**Figure 2.**
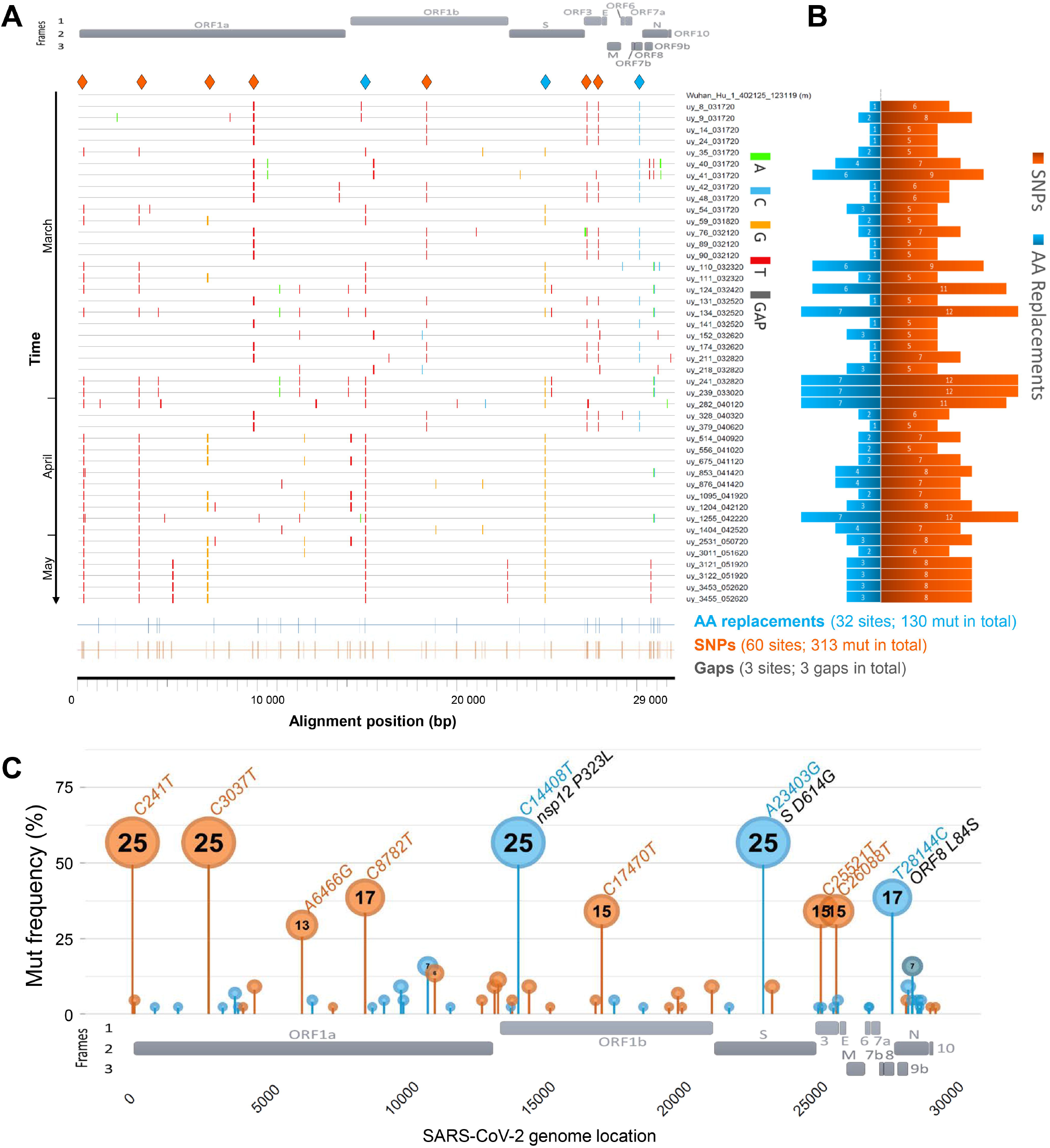
SARS-CoV-2 mutation patterns over time and mutation hotspots along the genome. **(A)** Highlighter plot showing mutations (mut) of Uruguayan study sequences compared to the reference Wuhan.Hu.1 sequence as master (on top). Mutations are shown as ticks, color-coded according to the legend to the right. Study sequences are sorted along the y-axis according to sampling time, with the earliest sequences on top and most recent sequences at the bottom. A SARS-CoV-2 genome map with the three reading frames’ coding genes is shown for orientation on top. All single-nucleotide polymorphisms (SNPs) are summarized in orange, and all SNPs resulting in amino acid (AA) replacements are summarized in blue at the bottom of the plot at the respective alignment positions. SNPs that are prevalent in >30% of study sequences are highlighted by orange or blue (if AA replacement) diamonds on top of the plot. **(B)** Mirror bar chart summarizing the number of SNPs (orange) and AA replacements (blue) per study sample, aligned with the study sample IDs in A. (C) Lollipop plot summarizing the frequency of SARS-CoV-2 mutations in the Uruguayan study cohort (n=44), using the same color code as in B. A SARS-CoV-2 genome map with base-pair positions is shown at the bottom. The bubbles’ y-coordinates indicate mutation frequencies, which are also shown inside the bubbles for mutations with >10% prevalence. Mutation details are shown in orange (SNPs, bp mutation) or blue and black (AA replacements, bp mutation and aa mutation/affected protein region) for mutations >30% prevalence.

### Phylogenetic assessment of the regional SARS-CoV-2 outbreak

To determine phylogenetic and epidemiologic characteristics of the Uruguayan SARS-CoV-2 outbreak, we performed a comprehensive set of phylogenetic analyses, including maximum-likelihood trees, haplotype networks, Nextstrain-based phylogenetic placements, and Bayesian phylogeographic analyses using BEAST (**Figures 3 and 4, Figures S5-S8**). Focusing on all available Uruguayan SARS-CoV-2 sequences that passed our internal and GISAID’s quality assessment (n=73), we observed an intermixing of our (44) and other (29) Uruguayan study sequences, both in maximum-likelihood IQ trees and in genetic-distance-based haplotype networks (**Figure S5**). Consistent with the mutational analysis of our internal data set (**Figure 2**), the phylogenetic analysis of the publicly available Uruguayan sequences reveals a predominance of clades S and G (G, GH, and GR), the former occupying most of the upper half of the maximum-likelihood tree (**Figure S5A**) and the right half of the haplotype network (**Figure S5B**), the latter the respective opposite halves, highlighted by green and red arrows for spike D614G and ORF8 L84S key mutations, respectively. The allocation of neighborhood data on the phylogenetic tree indicated regionally clustered appearances of phylogenetically related viruses, as most evident for Carrasco, Pocitos, Malvin, Reducto (all in Montevideo), and Rivera (**Figure S5A**).

**Figure 3.**
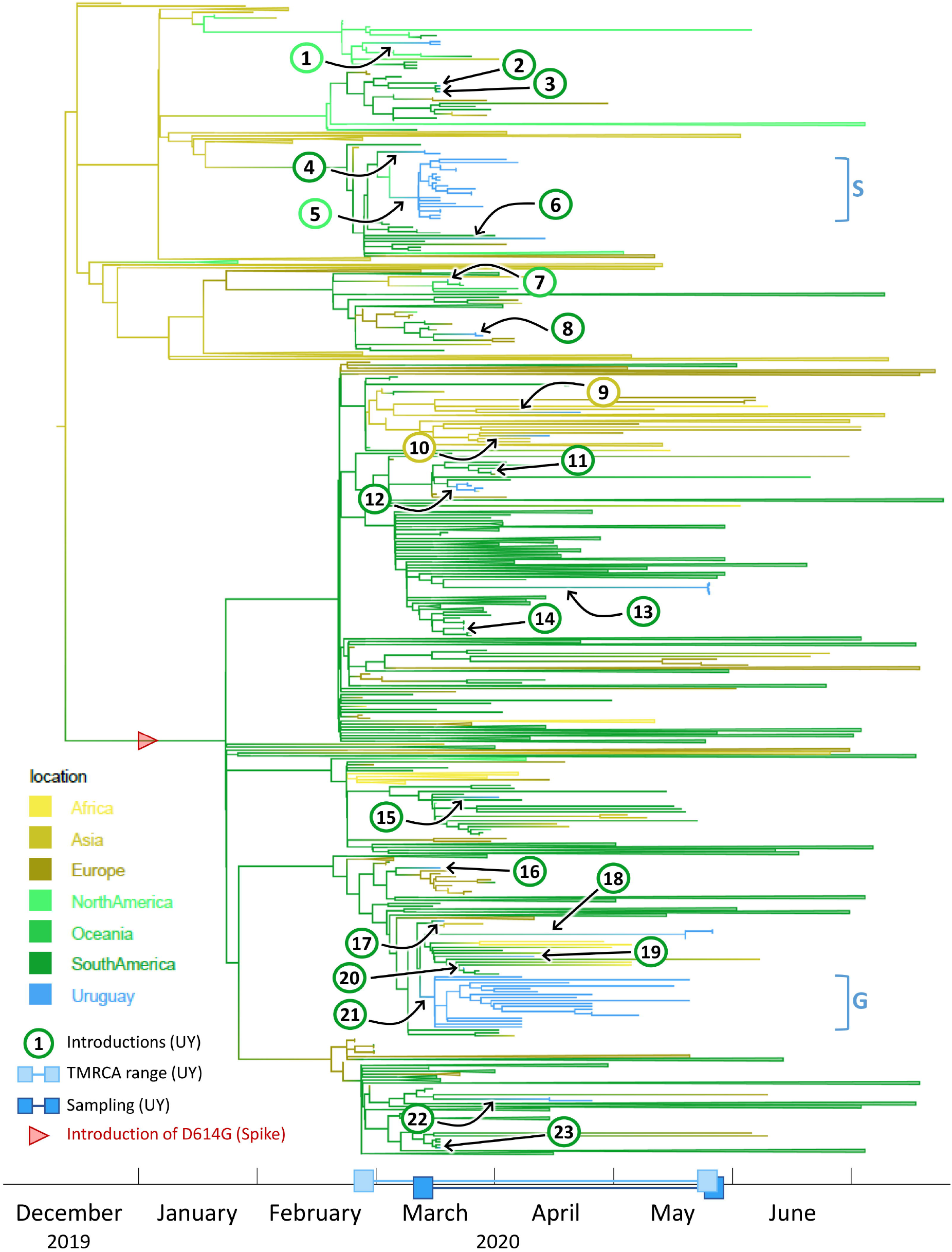
Time-scaled phylogenetic tree to identify the source regions of the sequences in the imported Uruguayan clusters. We employed a discrete state phylogeography diffusion model in BEAST with seven ancestral location states (Africa, Asia, Europe, North America, Oceania, South America, and Uruguay) to identify the most probable source locations for the sequences in the 23 previously identified international introductions into Uruguay. The branches of the trees are color-coded according to each geographic region. A color gradient along the branches indicates historic introduction events between locations. The introductions into Uruguay are highlighted by black arrows and circles with consecutive numbering according to the introduction event (color-code of circle outline: probable source continent). The times of most recent common ancestors (TMRCAs) of Uruguayan (UY) sequences and their sampling period are indicated as ranges along the x-axis timeline. Branches that are not involved in introduction events are collapsed to facilitate visualization. The introduction of the spike D614G mutation is indicated by a red arrowhead. The two major Uruguayan clades are highlighted by brackets, and GISAID clades are indicated.

**Figure 4.**
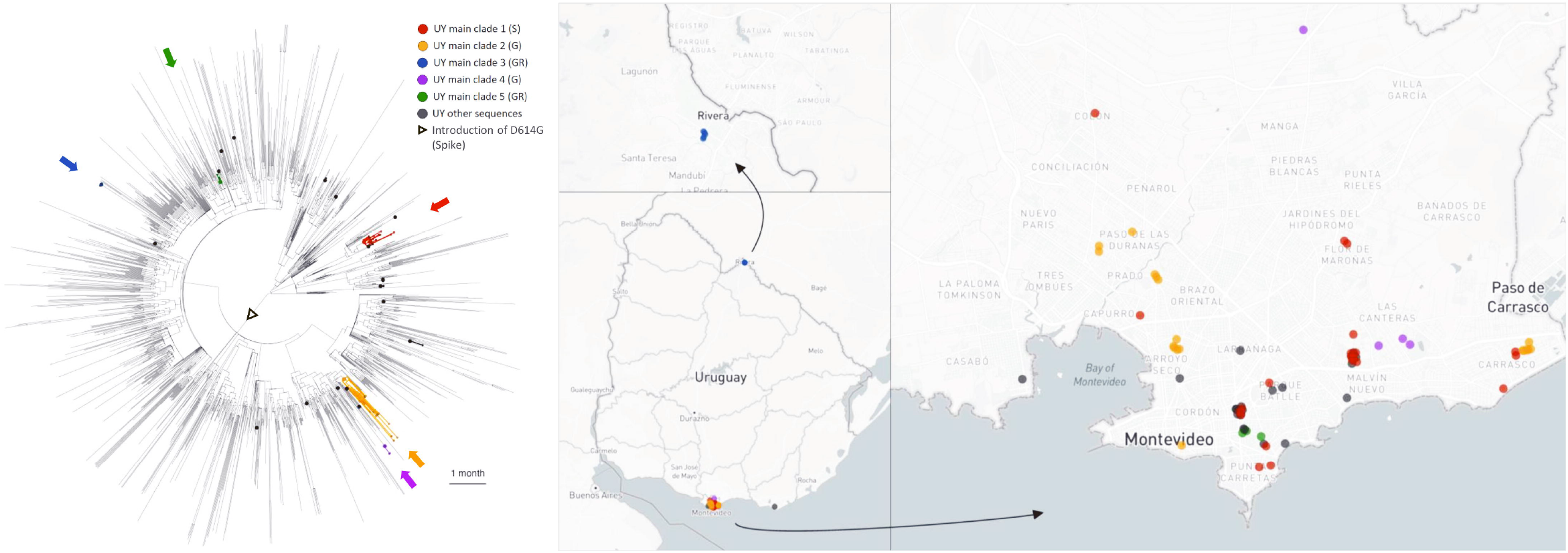
Visualization of the evolutionary relationships and spatial distribution of SARS-CoV-2 samples in the five Uruguayan clusters. A time-scaled maximum clade credibility tree (MCC) was generated by the discrete phylogeographic analysis of 1810 SARS-CoV-2 genomic sequences. Uruguayan sequences are shown as colored circles, both in the phylogenetic tree and in the Uruguayan maps. The five main Uruguayan clusters are color-coded according to the legend (clades indicated in brackets). The remaining Uruguayan sequences, which are based on introduction events that did not form subsequent transmission chains within Uruguay, are shown as gray circles.

### Phylogeographic BEAST analysis reveals 23 introductions into Uruguay mostly from surrounding South American countries, resulting in five clusters

To assess global introductions and the regional spread of SARS-CoV-2 in Uruguay, we performed discrete phylogeographic analysis using BEAST with all 73 Uruguayan sequences, complemented with 1737 global reference sequences, based on the global subsampling dataset suggested by Nextstrain (**Table S2**). Genetic distance-based haplotype networks indicated genetic relationships of Uruguayan sequences with those from other South American countries, Europe and Asia (**Figure S6**).

By explicitly considering the time and sampling locations of our sequences, BEAST analyses revealed further important details about the evolutionary relationships of our sequences. The time-scaled maximum clade credibility tree (MCC) generated by the discrete phylogeographic analysis of our 1810 SARS-CoV-2 sequences is presented in **Figure S7** and shows a minimum of 23 independent introductions of the SARS-CoV-2 virus occurred. Collectively, these introduction events included representatives from seven of the GISAID clades (G, GH, GR, L, O, S and V) circulating worldwide and all were imported to the capital city Montevideo with the exception of one imported to Rivera.

The results of the discrete phylogeography analysis we performed that considered seven ancestral state locations further revealed that 18 of the 23 independent introductions are inferred to have originated from other South American countries. The remainder include two independent introductions of GISAID clade GR viruses from Asia, two from North America, and one clade V virus from Oceania (**Figure 3**). The viral sequences from the only two participants in our cohort that reported a possibility of travel-related infection after returning from Japan are inferred to have been imported from Asia and grouped on the tree within separate clades of Asian sequences. The estimated time to the most recent common ancestor (TMRCA), represented by the age of the root node of the entire tree, is in mid-December 2019 (9^th^-21^st^ of December), which is in agreement with the estimated origin of the pandemic in the Hubei Province in China, sometime between October and December, and the first contracted case in China recorded in mid-November [28-31].

Only five of the 23 independent virus introductions into Uruguay resulted in monophyletic clades with more than two sampled sequences in the country. These clades comprised 21, 18, 5, 4, and 3 sequences, respectively (**Figures 3 and 4, Figure S8**), suggesting that these viral outbreaks were maintained by community transmission once introduced into Montevideo.

To investigate the timing of the introduction of the viruses that founded these five main clades circulating in Montevideo, we estimated the time of their most recent common ancestor (TMRCA), acknowledging that the actual introduction events likely occurred even before the corresponding TMRCAs.

The TMRCA of the first main clade of 21 sampled sequences, highlighted in red in **Figure 4**, was estimated to fall between the 2^nd^ and 5^th^ of March, 2020 and involved the importation of a GISAID clade S virus. Viruses within this transmission cluster were restricted to Montevideo, where they were distributed among nine of the local neighborhoods, including two Hospitals and one research institute (**Figure S8**). A North American sequence from Mexico was positioned basally to this clade on the tree identifying this country as the most plausible source location.

The TMRCA of the second main clade, which comprised of 18 virus sequences, highlighted in yellow in **Figure 4**, was estimated to fall between the 13^th^ and 17^th^ March 2020 and involved the import of a GISAID clade G virus. Viruses within this transmission cluster were restricted to the city of Montevideo, where they were distributed among nine neighborhoods and included samples from two nursing homes and one hospital (**Figure S8**). The viral sequences in this clade were inferred to have a South American origin.

The TMRCA of the third main clade of five sampled sequences, highlighted in blue in **Figure 4** and corresponding to a GISAID clade GR, was estimated to fall between March 20^th^ and May 26th, 2020. This transmission cluster consisted of five viruses from the Hospital de Rivera in Rivera, a small city situated on the border with Brazil (**Figure S8**). This clade was inferred to have originated in South America and groups with a sequence from Brazil on the MCC tree suggesting this was the source location for this viral introduction

The TMRCA of the fourth main clade with four sampled sequences, highlighted in purple in **Figure 4**, was estimated to fall between March 14^th^ and May 20^th^, 2020 and involved the introduction of a GISAID clade G virus into Montevideo (**Figure 4**). This clade comprised four viruses from within two Montevideo neighborhoods (**Figure S8**). The virus responsible for this introduction was also inferred to have originated in South America.

The TMRCA of the fifth main clade of three sequences, highlighted in green in **Figure 4**, was estimated to have occurred between 22^nd^ and the 24^th^ March 2019 and involved the introduction of a GISAID clade GR virus into Montevideo. This clade comprised three viruses from within a single Montevideo neighborhood (**Figure S8**). Collectively, sequences from the four main clades described above were sampled between March and May 2020 and were distributed among 17 of the 57 neighborhoods or *barrios* in Montevideo.

All of these five independent introduction events that formed sustained transmission chains identified here were estimated to have occurred close to the time of the first officially diagnosed case in Uruguay on March 13^th^, 2020 and the putative date of origin of the pandemic in Uruguay on March 7^th^, which was thought to be introduced by a single female traveler who arrived in Montevideo on a flight from Italy and subsequently attended a wedding reception in the city that was attended by over 500 guests.

### Distribution of SARS-CoV-2 within Uruguay

Within these five clades we identified a total of ten sequence clusters (sequences from the same institution that group together on the tree), spread across six of the seven health institutions from which we had more than one sample (**Figure S8**). These included one cluster of five GISAID clade GR sequences in the Hospital de Rivera (red dots), two clusters of two clade G sequences in the Hospital Vilardebó (green dots), three clusters each comprised of two clade S sequences in the Institut Pasteur (brown dots), and two clusters from the Asociación Española Primera en Salud (blue dots) comprising two S and two G clade sequences (**Figure S8**).

### Clinical correlations separate from mutational correlation clusters

The availability of study participants’ clinical and demographic data combined with mutational data of the infecting SARS-CoV-2 viruses enabled us to perform comprehensive correlation analyses (**Figures 5 and 6, Figures S9-S11**). Overall, Spearman rank correlation analyses revealed six prominent correlation clusters of significant positive or mixed correlations (**Figure 5, Figure S9**). Four of the highlighted clusters in **Figure 5** (clusters 1-4) are dominated by mutations that form mixed clusters with demographic parameters such as sampling location or treating healthcare institution and a few other parameters. Clusters 5 and 6 are different in forming separate clinical correlation clusters together with the parameters “sex” (cluster 5) or “age” (cluster 6) without essential associations to virus mutations. Cluster 5 reveals a significant association of female sex with clinically asymptomatic courses of the disease and a lower risk of developing fever. Cluster 6 indicates a network of positive correlations among age, lethal outcome, and five clinical parameters. In addition to the highlighted clusters, we observed some smaller clusters, mainly composed of mutations. The tight network of positive clinical correlations and a more outspread correlation network of clades with regional appearances and selected demographic parameters are shown in greater detail in **Figure 6A**. Specifically, statistical analyses of lethal outcome as study parameter revealed significant positive correlations with arterial hypertension (AHT), kidney failure and ICU admission complemented by borderline-significant associations with additional clinical parameters (hospitalization, diabetes mellitus II, and obesity) and age, but no association with any specific mutation (**Figure 6B, Figure S10**). Accordingly, the spike D614G mutation and clade G-related viruses, in consequence, are not associated with any clinical parameters, severity or lethality. D614G only correlates with co-occurring/inversely occurring mutations, treating healthcare institutions, and time since sampling started (**Figure 6C, Figure S11**). Fatality rates among clade G, GR, and S-infected individuals were comparable at 18% (3/17), 14% (1/7), and 12% (2/17), respectively.

**Figure 5.**
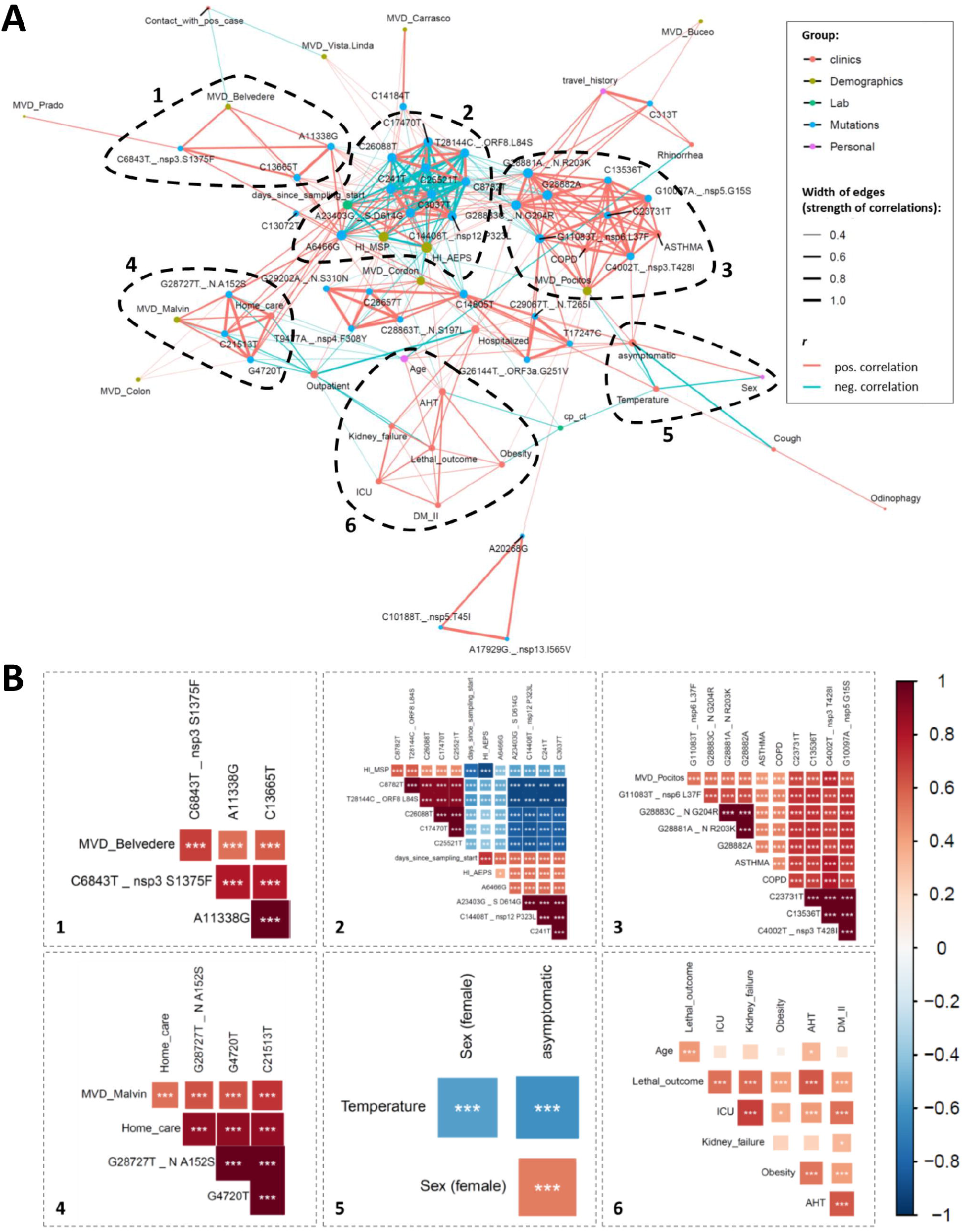
Correlation network analysis of virologic, demographic, and clinical parameters among Uruguayan study samples/participants. **(A)** In the correlation network plot, nodes represent clinical, demographic, laboratory, mutational, and personal parameters, and red and blue edges represent positive and negative correlations between connected parameters, respectively. Only significant correlations (*P*<0.05) are displayed between parameters with at least two positive events. Edge width corresponds to the strength of the correlation. Nodes are color-coded based on the grouping in clinical, demographic, laboratory, mutation, or personal parameters according to the legend to the upper right, and node size corresponds to the degree of relatedness of correlations. The six most prominent mixed correlation clusters are encircled with dashed lines and shown in greater detail as correlograms in the dashed boxes with matching numbers (1-6). **(B)** In the correlograms, squares are sized and color-coded according to the magnitude of the correlation coefficient (*r*). The color code of *r* values is shown to the right (red colors represent positive, blue colors negative correlations between two parameters). Asterisks indicate statistically significant correlations (*P < 0.05, **P < 0.01, ***P < 0.005). Correlation analysis was done using nonparametric Spearman rank tests. MVD: Montevideo, HI: healthcare institution, ICU: intensive care unit, AHT: arterial hypertension, DM II: diabetes mellitus type II, COPD: chronic obstructive pulmonary disease.

**Figure 6.**
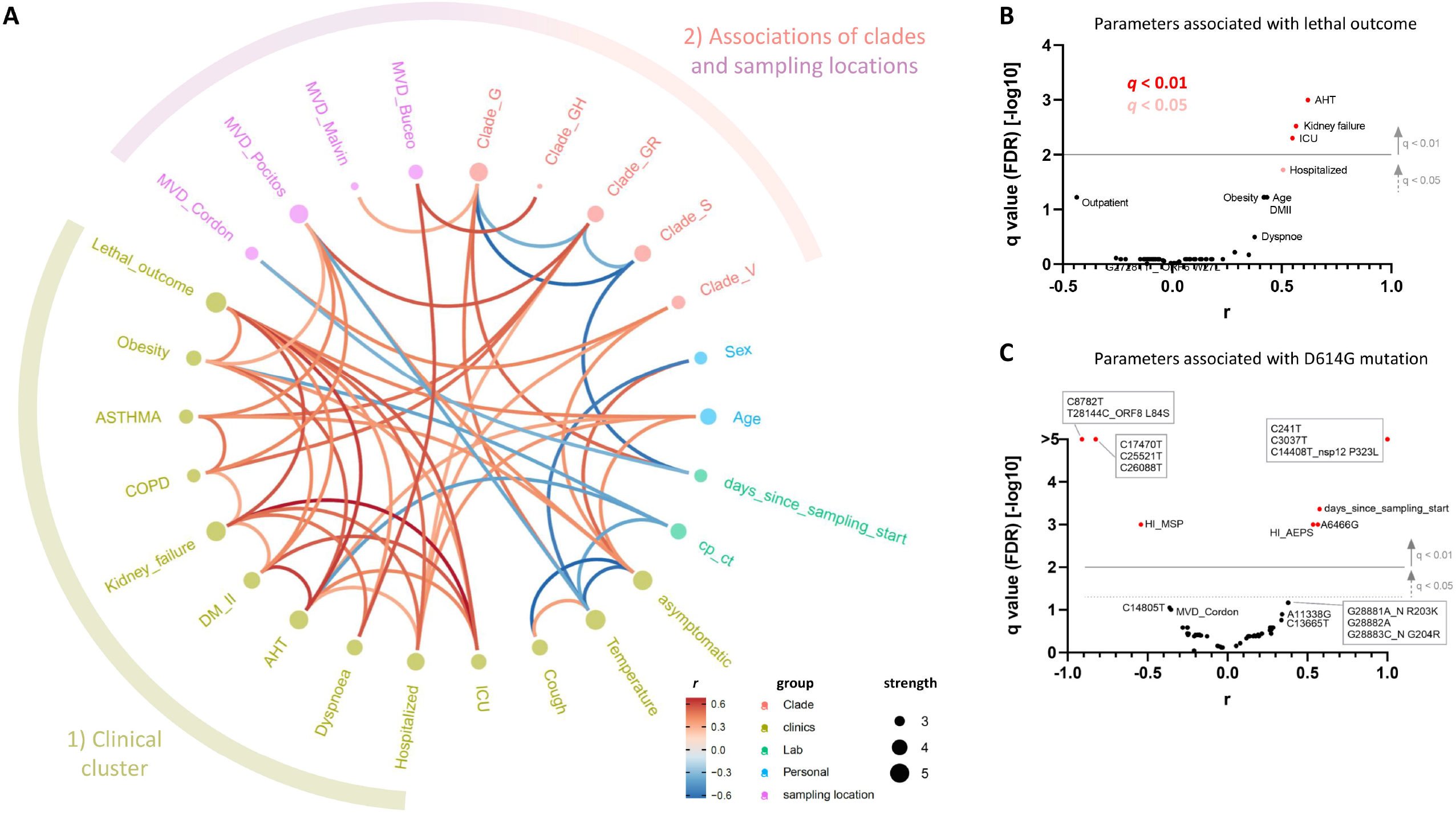
Clade and clinical correlation analysis and study parameters associated with lethal outcome and spike D614G mutation. **(A)** In the edge bundling correlation plot, red and blue edges represent positive and negative correlations between connected parameters, respectively. Only significant correlations (*P*<0.05) are displayed, and all parameters have at least two positive events. Nodes are color-coded based on the grouping into clades, clinical, laboratory, neighborhood, and personal data according to the legend at the bottom. Node size corresponds to the degree of relatedness of correlations. Surrounding circle segments highlight a strong clinical cluster and a less pronounced clade/sampling location cluster. **(B)** Volcano plot of parameters associated with lethal outcome. The full data set (see **Figure 5** and **Figure S9**) was screened for parameters with false discovery rates (FDR) of *q*<0.01 (red, considered significant) and 0.01<*q*<0.05 (pale red, considered borderline significant). **(C)** Volcano plot of parameters associated with the presence of D614G mutation in spike proteins of study participants’ infecting SARS-CoV-2 viruses. The same display was used as in B. Correlation analysis was done using nonparametric Spearman rank tests. All parameters that achieved *P* < 0.05 correlations are labeled. Parameters inheriting the same dot are boxed. MVD: Montevideo, HI: healthcare institution.

## Discussion

The Uruguayan epidemic is characterized by an early clade S dominance that was subsequently replaced by clade G variants (**Figures 1 and 2**), which matches the global trend [27,32,33]. Uruguayan clade S viruses are characterized by the key mutations T28144C, causing the AA replacement L84S in ORF8, and C8782T, in agreement with global clade S strains [27]. In our data set, we observed three additional co-occurring SNPs in ORF1b and ORF3 that were present in 15 out of 17 clade S variants, i.e., C17470T, C25521T, and C26088T. These mutations are less common and are presumably characteristic of the regional outbreak. Further studies need to show whether this subclade will be fixed in regional and/or supraregional epidemics, and whether founder effects or functional features accounted for the early clade S dominance over the original clade as well as the subsequent fluctuating prevalence and decline. According to the CoV-GLUE database, ORF8 L84S is the 8th most prevalent AA replacement to date [34]. After a controversial debate about the ancestry of L and S types and the functional impact of L84S [35-38], the still limited amount of data indicates that L84S might confer selection advantage and render the virus more virulent based on destabilizing the immuno- and replication-modulatory protein ORF8 and mitigating binding of ORF8 to human complement C3b [35,37-39].

End of March/early April 2020, we observed a subsequent switch in dominance from clade S to clade G-variants (G, GR, and GH) (**Figures 1 and 2**). It positioned Uruguay somewhere in the middle in the asynchronous transition process from spike D614 to G614 virus predominance, i.e., between the early European and the mostly late Asian countries [32,33]. Diverse structural and functional assays strongly suggest that the spike D614G mutation renders SARS-CoV-2 more infectious by stabilizing its structure, i.e., through impact on the, compared to SARS-CoV-1, even more fragile metastable SARS-CoV-2 spike protein, thus reducing the shedding of the S1 subunit. Furthermore, D614G triggers higher spike numbers on the virion surface and induces a more open, receptor-binding domain (RBD)-up spike conformation toward a receptor-binding and fusion-competent state [33,35,37,40-44]. In D614G spikes, binding to the angiotensin-converting enzyme 2 (ACE2) receptor is not increased, and SARS-CoV-2 viruses do not acquire D614G escape mutations *in vitro* under neutralizing antibody immune pressure [40,45]. Instead, D614G increases neutralization susceptibility of SARS-CoV-2, which assures high sensitivity to vaccination-induced neutralizing antibodies [46,47].

Although D614G serves as the clade G-defining mutation with likely effects on virus infectivity/transmissibility, D614G is governed by a very strict co-appearance with C241T, C3037T, and C14408T, both in Uruguayan and global G variants (**Figure 5, Figure S5**) [27]. Notably, in addition to the D614G-causing A23403G mutation in spike, C14408T is responsible for the AA replacement P323L in the RdRp gene. Based on their central roles in viral entry and replication, their co-evolution is of particular interest, and a mutual contribution to the selective advantage of G-haplotypes is assumed [33]. Interestingly, none of the viruses harboring a single clade G mutation prevailed to achieve epidemiological relevance, e.g., D614G alone or P323L alone have ≤0.3 global prevalence, whereas D614G and P323L together have ∼70% global prevalence as of August 2020 [33]. P323L, although not located in the active center, possibly influences RdRp fidelity through allosteric effects at the nsp12 interface with the nsp8 cofactor and might increase the viral mutation rate [33,39,48]. Thus, the strong correlation patterns between key mutations in our Uruguayan data set, mirroring global patterns, allows us hypothesize that coupled mutations, such as D614G in spike and P3233L in RdRp might synergize for the epidemiological success of the virus. It may allow a fine balance between efficient transmission (e.g., by D614G, even in asymptomatic cases) and limited clinical presentation (e.g., by P323L, decreasing the production of viral RNA) to eventually attenuate an aggressive virus that, as shown for MERS and SARS-CoV-1, is more vulnerable to viral clearance with lack of long-term epidemiological success. Further studies are needed to determine how the increasing diversity of mutation patterns influence the fitness and reproduction of viral populations, the susceptibility/evasion to immune responses and treatment, and how these mutations are selected in the human body or during transmission.

Beyond the functional relevance of emerging and transmitted mutations, they serve as a fine tool to dissect population phylodynamics, transmission chains and epidemiologic clusters. The number of independent introduction events into a particular country as a proportion of the total number of sequences in the data set is considered a rough measure of the relative influence of intercontinental and international travel on the subsequent epidemiological dynamics within that country. For Uruguay, with 23 identified independent migration events out of a total of 73 viral sequences in our dataset, this proportion is relatively low if compared to other countries such as Belgium (331/740) [11] but higher than others like Brazil (>100/490) [8]. It possibly indicates that the relative influence of intercontinental and international travel has been less important in driving the dispersal dynamics of the Uruguay outbreak compared to other, larger or more connected countries like Belgium. The town of Rivera, situated on the border with Brazil, has been a main concern for the government and many outbreaks occurred along the border that were rapidly brought under control. In line with our BEAST data that suggested a single transmission from Brazil as the main source of the Rivera infections, the first Rivera outbreak was reported to have begun when a COVID-19-positive Uruguayan was diagnosed on May 7^th^, 2020. Our BEAST analysis strongly supports the former hypothesis from the Uruguayan Government and Ministry of Health (MSP) that a local metallurgic worker that used to travel to Brazil everyday had been infected with SARS-CoV-2 in Brazil and introduced this strain to Rivera [49,50].

There are two main reasons for why the extent of the geographical distribution and the density of viruses in each neighborhood within Montevideo and Rivera responsible for the five main clades of SARS-CoV-2 circulating in Uruguay (**Figure 4, Figure S8**) represent an underestimate of the true values of both these variables. First, our 73 Uruguayan sequenced viral genomes represent only a relatively small fraction of the total number of infections that occurred in the actual outbreak seeded by these viruses, with estimates obtained from contact tracing efforts suggesting that collectively these viruses infected at least 364 individuals [2]. Secondly, 39 (53%) of our Uruguayan samples were either collected in hospitals (Hospital de Rivera, Asociacion Española - AEPS, or Hospital Vilardebó) where the infected patient was treated or where the infection was acquired, in nursing homes, or in research institutes (Institut Pasteur) where samples were processed. In these cases, the home address of the patient samples is not publicly available. This precluded the adoption of the continuous diffusion phylogeography model [51] that makes use of the actual geographic coordinates of the samples to infer the transmission links among the sampled locations in the various Montevideo neighborhoods, and instead, limited our analysis to describing the spatial relationships among the sampled SARS-CoV-2 sequences.

Our discrete phylogeographic analysis provided a new perspective to the believed origin of the Uruguayan SARS-CoV-2 outbreak from overseas by a single traveler returning from Europe [52-54]. While overall similarities of Uruguayan sequences with European and Asian sequences were observed in mutation patterns and genetic distance-based haplotype networks, our phylogeographic analyses using BEAST indicate that Uruguayan introductions that resulted in outbreaks were mostly restricted to neighboring South American countries (**Figure 3**), which stands in contrast to the suggestions of a recent preprint article [7]. Instead, our data support the idea that the outbreaks that were seeded after return from overseas travel, were contained successfully through social distancing, mask wearing, rigorous testing, contact tracing, and partial lockdown [2].

Having determined the regional SARS-CoV-2 mutation patterns and phylogeographic spread, the question remained whether and to what extent genomic features are coupled to demographic and/or clinical parameters. Our correlation data revealed significant clusters of co-occurring or mutually excluding mutations with regionally accumulated appearances of clades/mutation clusters (**Figures 4 and 5, Figure S5**). In contrast, the large bulk of clinical parameters clustered separately without major influence from viral mutations (**Figures 5 and 6, Figure S10**), which is in line with a recent publication that reported no significant impact of genetic variation on clinical outcome [55]. More specifically, we studied pairwise correlations with the spike D614G mutation, which, because of perfectly matching mutation patterns, also represents correlation analyses for the RdRp P323L mutation or G-related clades (G, GR, and GH). In an FDR-adjusted analysis, the presence of D614G mutation was coupled to other viral mutations, treatment of the infected patients in a regional healthcare institution, and late sampling, but not to clinical parameters (**Figures 5 and 6, Figure S11**). There have been controversial reports of D614G being associated with higher fatality rates and/or severe illness in a few data sets [56,57], whereas more recent data suggests no correlations of D614G with clinical outcome [33], the latter supporting our findings. Our analysis of associations with clinical parameters pointed at significant associations between lethal outcome and arterial hypertension, kidney failure, and ICU admission, which mirrors clinical studies on associated factors or predictors of disease severity/progression [58,59].

In sum, our characterization of Uruguayan SARS-CoV-2 phylogenetics, mutation patterns, and their correlation with demographic and clinical parameters did not identify critical viral attenuations or clinical peculiarities that can primarily account for the exceptionally well curbed regional COVID-19 epidemic [2]. It instead suggests that socio-epidemiologic mitigation strategies managed to curtail COVID-19 to restricted regional transmission clusters in Uruguay.

We hope that these findings contribute to define the South American COVID-19 outbreak better, to optimize and develop efficient, fast, and low-cost mitigation strategies and diagnostic pipelines for Uruguay and other countries, and to assist physicians dealing with strategies for this and future emerging infections.

## Data Availability

All data referred to in the manuscript are fully available without restriction, and a summary is provided in Table S1 of the manuscript. All SARS-CoV-2 sequences are available from the GISAID database (accession numbers listed in Table S1).

## Acknowledgments

The authors thank all participants who agreed to participate in this study and the healthcare personnel in Uruguay for their dedication to COVID-19 patients’ care. We wish to acknowledge the support of New York University’s Data Services, Bobst Library, and, in particular, the expertise shared by Christopher Schwarz and Senior Academic Technology Specialist Denis Rubin in software script development. We also acknowledge the support of the NYU Langone Health High-Performance Computing resource and the Laura and Isaac Perlmutter Cancer Center, which partially supports the Genome Technology Center. We thank the scientists across the world who deposited SARS-CoV-2 sequences in GISAID, especially those who deposited 29 Uruguayan sequences that supplemented our 44 sequences. We would also like to thank Flavia Camacho for administrative assistance.

## Contributors

VE1, AH, and RD conceived research goals, experiments, and analyses. GWH, BM, SS, VP, and RD performed formal analyses. VE1, RB, and AH acquired funding for the project. PZ, CM, VP, NM, CB, SI, MF, VP, VE2, GM, AS, FR, MN, LP, SB, and MNZ performed experiments and/or were involved in sample acquisition. BM, MM, GWH, SD, AH, and RD developed methodologies, designed, and/or implemented computer codes. VE1, GWH, MM, SD, RB, AH, and RD supervised the research activity and validated the research output. GWH, BM, SD, VP, and RD prepared figures and tables. VE1, GWH, AH, and RD wrote the manuscript. All authors reviewed and edited the manuscript.

## Supporting Information

**Table S1. Study samples, demographic, and clinical parameters of COVID-19 participants**.

Abbreviations: GF: GeneFinder, na: not available, NOS_C: naso- and oropharyngeal swabs combination, TIB: TIB ROCHE, TS: tracheal secretions. hCoV-19/Uruguay/UY-NYUMC932/2020|EPI_ISL_480430|2020-04-14 was removed from analyses because of > 35% undefined base pairs (N’s) in full genome sequence.

**Table S2. Origin and number of global study sequences used for Bayesian analyses**.

**Figure S1. Map of SARS-CoV-2 sample collection in our Uruguayan study cohort**.

Map of Montevideo, Uruguay, with sampling locations indicated by dots that are colored by sample numbers according to the legend to the right. Sampling density was most significant around the center of Montevideo, indicated by a gray-to-red density gradient.

**Figure S2. SARS-CoV-2 amino acid replacements over time**.

Highlighter plot showing amino acid (AA) replacements of Uruguayan study sequences compared to the reference Wuhan.Hu.1 sequence as master (on top). Mutations are shown as ticks, color-coded according to the legend at the bottom. Study sequences are sorted along the y-axis according to sampling time, with the earliest sequences on top and most recent sequences at the bottom. A SARS-CoV-2 genome map is shown on top with protein regions consecutively assembled as done for the AA alignment. The gray tones of the protein bars relate to the reading frames of their coding genes. All occurring AA replacements are summarized at the bottom of the plot (compressed). AA replacements that are prevalent in >30% of study sequences are indicated by diamonds on top of the plot in the color of the replacing amino acid. A bar chart is shown to the right summarizing the number of AA replacements per study sample, aligned with the highlighter results and sample IDs on the left.

**Figure S3. Summary of SARS-CoV-2 BP and AA mutations in the cohort and per subject**.

**Left**: Bar diagrams showing mutation counts per site in the entire cohort. All single nucleotide polymorphisms (SNPs) are shown on the upper left, with total counts indicated on the y-axis and as numerical values on top of each bar. The base pair mutations are indicated on the x-axis together with the amino acid (AA) replacement, if applicable. On the lower left, all AA replacements are shown separately. **Right**: Bar diagrams showing cohort-wide mutation counts per genome. SNPs are shown on the upper right, AA replacements on the lower right. The number of mutations per genome is listed on the x-axis, and the occurrences in the cohort on the y-axis and as numerical values on top of each bar.

**Figure S4. Uruguayan SARS-CoV-2 mutation clusters**.

Correlogram of associations among mutations observed in our Uruguayan study cohort (n=44) with squares sized and color-coded according to the magnitude of the correlation coefficient (*r*). The color code of *r* values is shown to the right; red colors represent positive, blue colors negative correlations between two connected parameters on the x- and y-axes. Asterisks indicate statistically significant correlations (*P < 0.05, **P < 0.01, ***P < 0.005). The correlogram is shown with hierarchical clustering according to the dendrogram at the bottom. The color-strip indicates gene relatedness of mutations according to the color code in the legend. Correlation analysis was done using non-parametric Spearman rank tests.

**Figure S5. Phylogenetic and mutation network analysis of Uruguayan SARS-CoV-2 viruses**.

**A**. Maximum-likelihood IQ-tree of 73 Uruguayan SARS-CoV-2 sequences and Wuhan.Hu.1 reference sequence, run with 1000 Bootstrap replications. Branch symbols and taxa (ID colored ranges) are colored according to country and study source, as explained in the figure legend. The introductions of key mutations are shown by red and green triangles. For each sample, reported/estimated site of infection/sampling location (est: estimated, hos: treating hospital), clades, and time since global outbreak are indicated by the circular color strip around the tree according to the legend. Clustered appearances of clades at specific sites/neighborhoods are highlighted by arrows and labeled. **B**. Genetic distance-based haplotype network analysis of Uruguayan SARS-CoV-2 sequences. Circles represent populations of sequences with identical mutation patterns (haplotypes) as compared to Wuhan.Hu.1 as reference (gaps and missing data not considered). The circles are sized and colored relative to the number and source countries of contributing sequences, respectively. One sample ID annotates each haplotype population representatively. Ticks on the connecting lines indicate discriminating mutations between haplotypes. Two major branch-defining mutations are highlighted as red and green ticks according to the legend. Red and green polygons encircle haplotype populations carrying respective key mutations (spike D614G and ORF8 L84S).

**Figure S6. Haplotype network analyses of Uruguayan SARS-CoV-2 sequences among global reference strains**.

Genetic distance-based haplotype network analysis of 73 Uruguayan SARS-CoV-2 sequences among 609 subsampled, global SARS-CoV-2 sequences. Circles represent populations of sequences with identical mutation patterns (haplotypes) as compared to Wuhan.Hu.1 as reference (gaps and missing data not considered). The circles are sized and colored relative to the number and source countries of contributing sequences, respectively. One sample ID annotates each haplotype population representatively. Ticks on the connecting lines indicate discriminating mutations between haplotypes. The node carrying the Wuhan-Hu-1 reference sequence is labeled, and haplotypes sharing the spike D614G mutation among the sequences of the reticular network are encircled by a dashed, dark red polygon. The location of major and minor clusters of Uruguayan sequences is highlighted by large and small blue arrows, respectively.

**Figure S7. Time-scaled phylogenetic tree to identify Uruguayan clusters**.

A cluster is defined as a phylogenetic clade corresponding to an independent introduction into Uruguay. Uruguayan sequence tree branches are colored light blue and non-Uruguayan sequence branches gray. The two main clusters are highlighted by brackets, and their GISAID clades are indicated. The introduction of the spike D614G mutation is indicated by a red arrowhead.

**Figure S8. A visualization of the evolutionary relationships and spatial distribution of our SARS-CoV-2 samples in hospitals and nursing homes in Montevideo**.

Time-scaled maximum clade credibility tree (MCC) generated by the discrete phylogeographic analysis of 1810 SARS-CoV-2 sequences. According to the legend, Uruguayan sequences are shown as colored circles both in the phylogenetic tree and in the Uruguayan maps, color-coded based on their relationship to hospitals and nursing homes. Samples that are not associated with a hospital or nursing home are shown as gray circles.

**Figure S9. Correlation analysis of viral, demographic, and clinical parameters of Uruguayan study samples/participants**.

Correlogram of associations among indicated parameters as present in our Uruguayan study cohort (n=44) with squares sized and color-coded according to the magnitude of the correlation coefficient (*r*). The color code of *r* values is shown to the right; red colors represent positive, blue colors negative correlations between two connected parameters on the x- and y-axes. Asterisks indicate statistically significant correlations (*P < 0.05, **P < 0.01, ***P < 0.005). The correlogram is shown with hierarchical clustering according to the dendrogram at the bottom. The color-strip indicates group relatedness of parameters according to the color code in the legend. Correlation analysis was done using nonparametric Spearman rank tests. MVD: Montevideo, HI: healthcare institution.

**Figure S10. Statistics of correlations with lethal outcome of SARS-CoV-2 infection in our Uruguayan study cohort**.

Correlation statistics between lethal outcome and virus mutations are shown on the **left**, and between lethal outcome and clinical and demographic parameters in the **middle**. A summary of significant correlations, according to *q*<0.01, is shown on the **upper right**. For each correlation, correlation values *r, P* values, adjusted *P* values, and false discovery rates (FDR) *q* values are displayed together with significant discovery assessment. The distribution of *P* values across the data set is illustrated in a dot plot of ranked *P* values in the **lower right**. All significant results are highlighted in red. MVD: Montevideo, HI: healthcare institution, ICU: intensive care unit, AHT: arterial hypertension, DM II: diabetes mellitus type II, COPD: chronic obstructive pulmonary disease.

**Figure S11. Statistics of correlations with D614G spike mutation in infecting viruses of our Uruguayan study cohort**.

Correlation statistics between the presence of D614G spike mutation and other SARS-CoV-2 mutations are shown on the **left**, and between D614G spike mutation and clinical and demographic parameters in the **middle**. A summary of significant correlations, according to *q*<0.01, is shown on the **upper right**. For each correlation, correlation values *r, P* values, adjusted *P* values, and false discovery rates (FDR) *q* values are displayed together with assessment of significant discovery. The distribution of *P* values across the data set is illustrated in a dot plot of ranked *P* values in the **lower right**. All significant results are highlighted in red. MVD: Montevideo, HI: healthcare institution, ICU: intensive care unit, AHT: arterial hypertension, DM II: diabetes mellitus type II, COPD: chronic obstructive pulmonary disease.

