## Supplemental Methods for "SARS-CoV-2 genomic characterization and clinical manifestation of the COVID-19 outbreak in Uruguay"

***Sample collection, RNA extraction and diagnostic RT-PCR***

Naso-oropharyngeal swabs (NOS), bronchoalveolar lavages (BAL), nasopharyngeal aspirates (NA) and tracheal aspirates (TA) were collected in viral transport media (VTM) (Centers for Disease Control and Prevention, CDC; VTM, SOP# DSR-052-03) from suspect cases with acute respiratory symptoms, travel history to affected countries, and/or contact with COVID-19 confirmed cases. Sampling was done between March 17^th^ and May 26^th^, 2020, and samples were referred to the laboratory for diagnosis.

RNA was extracted from 300 or 500 μl per sample (in VTM) both using the QIAsymphonyⓇ DSP Virus/Pathogen Mini or Midi kit (Qiagen), respectively, and confirmatory qualitative commercial RT-PCR kits were used for diagnosis and screening (depending on critical availability during the outbreak) (**Table 1** in the main text). The criteria to consider a case positive is specified in the commercial kits’ datasheets [1-3]. For the only kit without specified cut-off, i.e., the RealStar SARS-CoV-2 RT-PCR kit (Altona), our laboratory used a calculated cut-off threshold of 40. The TIB BIOMOL has a cycle cut-off threshold of 36 (for the E gene) and 39 (for the RdRp gene); the GeneFinder commercial kit has a unique cycle cut-off threshold of 40 for the three genes (N, E, and RdRp genes).

***Library preparation and sequencing***

For viral genome amplification, we used the Swift Normalase Amplicon Panel (SNAP) SARS-CoV-2 Panel (Swift Biosciences, Whole viral genome single tube NGS assay, cat# SN-5XCOV296). First, isolated total RNA was converted to first-strand cDNA by random priming using the Superscript IV first-strand synthesis system (Invitrogen, cat# 180901050). Random priming was performed as follows: 65˚C for 5 min, cooled on ice followed by addition of mix to run cDNA first-strand synthesis 23˚C 10 min, 50˚C for 30 min, 80˚C for 10 min, and 4˚C hold. Ten microliters of first-strand cDNA were inputted into the two-step PCR reaction. The first step incorporates tiled primer pairs that target and enrich for the entire 29.9 kb COVID-19 viral genome (NCBI Reference Sequence NC_045512.2) during multiplex PCR. Multiplex PCR was run as follows: 98˚C 30 sec, four repeating cycles of 98˚C/ 10 sec, 60˚C/5 min, 65˚C/1 min. Eighteen repeating cycles of 98˚C/10 sec, 64˚C/1 min. The multiplexed samples were cleaned with 1x volume room temperature Ampure XP beads (Beckman Coulter, cat# A63882) for 5 min at room temperature, washed with 80% ethanol twice, and beads were resuspended in 17.4 µl of TE buffer (part of the swift kit). The indexing reagent mix was added to the resuspended beads and run in the indexing PCR reaction (each sample contains a unique i5 Illumina index) at 37˚C/20mins, 98˚C/30secs, eight repeating cycles of 98 ˚C/10 sec, 60˚C/30 sec, 66˚C/1 min.

After indexing PCR was completed, samples were cleaned with 1x volume Ampure XP beads (5 min bind time, 80% ethanol wash x2) and eluted in 30 µl of TE buffer. The final library products were run on Agilent Tapestation 2200 with high sensitivity DNA screen tape to verify the amplicon size of about 450 bp; each library was quantified using QPCR using the Kapa-Roche Library quant kit (Illumina, cat# KK4824) on the Bio-Rad cfx384 real-time system. Samples were normalized, pooled, and run on the Illumina NovaSeq 6000 system on a 300 cycle flow cell. Run metrics were paired-end 150 cycles with dual indexing reads. Both positive and negative control samples were also run during library prep but not sequenced. The negative control sample was water, and the positive control sample consisted of SARS-CoV-2 genome (Twist Biosciences, cat# 102024) serially diluted to 100 viral copies mixed into 50 ng of Universal Human Reference RNA (Agilent, cat# 18091050).

***Sequence read processing***

Reads were demultiplexed with Illumina bcl2fastq v2.20 requiring a perfect match to indexing barcode sequences; Illumina sequencing adapters were trimmed with Trimmomatic v0.39 (Bolger et al. 2014), and primers were removed with primerclip. Reads were aligned using BWA v0.7.17 [4] to a custom index containing human genome reference (GRCh38/hg38), including unscaffolded contigs and alternate references plus the reference SARS-CoV-2 genome (NC_045512.2, wuhCor1). Presumed PCR duplicates were marked using samblaster v0.1.24 [5]. Only sequences with >23,000 bp unmasked sequences were analyzed. Variants were called across all samples using bcftools v1.9:

bcftools mpileup --redo-BAQ --adjust-MQ 50 --gap-frac 0.05 --max-depth 10000 --max-idepth 200000 --output-type u |

bcftools call --ploidy 1 --keep-alts --multiallelic-caller -f GQ

Raw pileups were filtered using

bcftools norm --check-ref w --output-type u |

bcftools filter -i "INFO/DP>=10 & QUAL>=10 & GQ>=99 & FORMAT/DP>=10" --SnpGap 3 --IndelGap 10 --set-GTs . --output-type u |

bcftools view -i 'GT="alt"' --trim-alt-alleles

Viral sequences were generated by applying VCF files to the reference sequence using `bcftools consensus` with -m to mask sites below 20x with Ns, and -m N to mask sites of ambiguous genotypes with N. The codes used in data processing are available [6].

***Phylogenetic analyses***

***IQ trees.*** Maximum likelihood trees were performed using the IQ-TREE XSEDE tool, multicore version 2.0.6, on the Cipres Science gateway [7]. The best substitution model was determined among 87 options using ModelFinder, as implemented in the Cipres gateway. According to the Bayesian Information Criterion (BIC), TIM+F+I was chosen as the best-fit substitution model. Support values were generated with 1000 bootstrap replicates and the ultrafast bootstrapping method. Phylogenetic trees were visualized in Interactive Tree Of Life (iTOL) v.5 [8] or FigTree v.1.4.3 [9].

***Haplotype network analysis*** Genetic distance-based haplotype networks were created using DnaSP6 [10] and Population Analysis with Reticulate Trees (PopART) software [11-13]. Briefly, aligned fasta files with unique ten character codes and capped invariable ends were loaded, and ambiguous characters were replaced with N’s. Nexus and phylip haplotype files were generated considering sites containing gaps and missing data, and invariant sites were removed. An output file with all haplotype and polymorphism information was saved to support the generation of trait tables. The phylip haplotype file was imported as alignment into PopART together with location and/or mutation trait files, and median-joining networks created (epsilon = 0) with manually set trait colors.

***Mapping the distribution SARS-CoV-2 clades within Uruguay***

We investigated each of the SARS-CoV-2 clades’ spatial distribution resulting from independent introduction events in Uruguay, as detected using BEAST. In particular, we focused on the capital city, Montevideo, for which we had the densest sampling of genomic sequences. We investigated the geographic spread of identified clades and the detection of phylogenetic clusters in hospitals or nursing homes and other research institutes within Uruguay.

***Software Scripts and Visualization***

Correlograms were generated using the corrplot and RColorBrewer packages in program R [14] and Rstudio [15] as described recently [16]. Dendrograms were calculated using the dendPlot function and hclust method, or as implemented in the heatmap/complexheatmap packages in R. Annotations were done using heatmap, complexheatmap, and ggplot2 packages. Correlation network diagrams were generated in undirected mode in R and RStudio using ggraph, igraph, tidyverse, and ggplot2 packages, with clustering based on the igraph layouts graphopt or dh. Edges are weighted according to *P*-values (inversely). Edges are only shown if *P* < 0.05, and nodes without edges were removed. Nodes are sized according to the *r*-values of the connecting edges. Edge bundling graphs were generated in undirected mode in R and RStudio using ggraph, igraph, tidyverse, and RColorBrewer packages. Edges are only shown if *P* < 0.05, and nodes are sized according to the connecting edges’ r values. For lollipop plots, sequence retrieval and data manipulations were performed using dplyr (tidyverse) and seqinr packages, followed by data visualization using ggplot2.

Volcano plots were generated in Prism. Mirror bar charts were created in Microsoft Excel 2016. Multi-categorical alluvial diagrams were generated using RawGraphs with automatic sorting and 0.5 link opacity [17]. Stream graphs were created in RawGraphs using Expand representation as offset and Basis spline interpolation. Geocoding were done with the ggmap library in R/RStudio. Geographical contour heat maps with point annotations were generated using the packages ggmap, ggplot2, and dplyr.

1.	Altona. RealStar® SARS-CoV-2 RT-PCR Kit RUO 2020. Available from: <https://altona-diagnostics.com/en/products/reagents-140/reagents/realstar-real-time-pcr-reagents/realstar-sars-cov-2-rt-pcr-kit-ruo.html>

2.	GeneFinder. GeneFinder™ COVID-19 Plus RealAmp Kit 2020. Available from: <https://www.fda.gov/media/137116/download>

3.	Roche. Molecular diagnostic solutions

from Roche - SARS-CoV-2 tests 2020. Available from: <https://www.roche.de/res/content/11630/sars-cov-2_tests_ubersichtsbroschure_en__1_.pdf>

4.	Li H, Durbin R. Fast and accurate short read alignment with Burrows-Wheeler transform. Bioinformatics. 2009 Jul 15;25(14):1754-60.

5.	Faust GG, Hall IM. SAMBLASTER: fast duplicate marking and structural variant read extraction. Bioinformatics. 2014 Sep 1;30(17):2503-5.

6.	NYU-sequencing-core. Codes used in viral sequence data processing 2020. Available from: <https://github.com/mauranolab/mapping/tree/master/dnase>

7.	Miller MA, Pfeiffer W, Schwartz T, editors. Creating the CIPRES Science Gateway for Inference of Large Phylogenetic Trees. Gateway Computing Environments Workshop; 2010; New Orleans, LA.

8.	Letunic I, Bork P. Interactive Tree Of Life (iTOL) v4: recent updates and new developments. Nucleic Acids Res. 2019 Jul 2;47(W1):W256-W259.

9.	Rambaut A. FigTree. 1.4. Edinburgh, UK: 2012.

10.	Rozas J, Ferrer-Mata A, Sanchez-DelBarrio JC, et al. DnaSP 6: DNA Sequence Polymorphism Analysis of Large Datasets. Mol Biol Evol. 2017;34(12):3299-3302.

11.	Bandelt HJ, Forster P, Rohl A. Median-joining networks for inferring intraspecific phylogenies. Mol Biol Evol. 1999 Jan;16(1):37-48.

12.	Leigh J, Bryant D. PopART: Full-feature software for haplotype network construction. Methods Ecol Evol. 2015;6(9):1110–1116.

13.	PopART. Population Analysis with Reticulate Trees (PopART) 2020. Available from: <http://popart.otago.ac.nz>

14.	Team RC. R: A language and environment for statistical computing and graphics. Vienna, Austria: 2013.

15.	RStudio. RStudio Team: a bundle of RStudio’s popular professional software for statistical data analysis, package management, and sharing data products. Boston, MA: RStudio, Inc.; 2015.

16.	Tuen M, Bimela JS, Banin AN, et al. Immune Correlates of Disease Progression in Linked HIV-1 Infection. Front Immunol. 2019;10:1062.

17.	Mauri M, Elli T, Caviglia G, et al., editors. RAWGraphs: A Visualisation Platform to Create Open Outputs. 12th Biannual Conference on Italian SIGCHI Chapter; 2017; Cagliari, Italy: Association for Computing Machinery, New York, NY.
